# Maternal obesity before pregnancy predicts offspring blood pressure at 18 years of age: A causal mediation analysis

**DOI:** 10.1101/2020.11.22.20236398

**Authors:** Nicole Brunton, Brenden Dufault, Allison Dart, Meghan B. Azad, Jonathan M McGavock

## Abstract

**Importance:** Hypertension is the second most common pediatric chronic disease in Westernized countries. Understanding the natural history of hypertension is key to identifying prevention strategies.

**Objective:** Examine the relationship between maternal pre-pregnancy body mass index (BMI) and offspring blood pressure at 18 years and the mediating role of growth throughout childhood and adolescence.

**Design, Setting, and Participants:** We performed multivariable regression and causal mediation analyses within 3217 mother - offspring pairs from the Avon Longitudinal Study of Parents and Children (ALSAPC) prospective birth cohort. Latent trajectory analysis (LTA) was used to quantify the mediating variable of offspring BMI from 7 to 18 years of age.

**Exposures:** The main exposure was maternal pre-pregnancy BMI. Analyses were adjusted for relevant confounders including maternal education, maternal blood pressure, and weeks gestation at delivery.

**Main Outcomes and Measures:** The main outcome was offspring blood pressure at 18 years of age categorized as normal (SBP < 120 mmHg or DBP < 80mmHg) or elevated (SBP ≥ 120 mmHg or DBP ≥ 80 mmHg) as per the 2017 American Academy of Pediatrics guidelines.

**Results:** At 18 years of age, among 3217 offspring, 676 (21%) were overweight or obese, 865 (27%) had elevated blood pressure, and 510 (16%) were hypertensive. LTA identified five distinct offspring BMI trajectories. Multivariate logistic regression revealed that for every 1 unit increase in maternal BMI the risk of elevated blood pressure at 18 years of age increased by 5% (aOR: 1.05, 95% CI: 1.03 – 1.07; p <0.001) and this effect was reduced after adjusting for offspring BMI trajectory (aOR: 1.03, 95% CI: 1.00 – 1.05; p = 0.017). Causal mediation analysis confirmed offspring BMI trajectory as a mediator accounting for 46% of the total effect of maternal BMI on elevated offspring blood pressure (aOR 1.22; 95% CI: 1.07-1.39).

**Conclusion and Relevance:** Maternal BMI prior to pregnancy is associated with an increased risk of elevated blood pressure in offspring at 18 years of age and is mediated, in part, by offspring BMI trajectory throughout childhood and adolescence.

## INTRODUCTION

Hypertension is one of the most common pediatric chronic diseases in Westernized countries.^1^ Elevated blood pressure in adolescence is a strong predictor of developing hypertension^2^ and cardiovascular disease as an adult. While obesity status, genetic propensity, and nutritional factors contribute to pediatric hypertension etiology, other early life factors including in utero exposures, may also play a role.^3^ Life course research, natural experiments, and pre-clinical studies have all shown modest to robust associations between exposure to maternal overweight or obesity in utero, and an elevated risk for cardiovascular disease related risk factors, including hypertension, later in life. ^4-9^ As over 50 million women live with overweight or obesity during pregnancy, it one of the most prevalent adverse metabolic disorders to which children are exposed in utero.^10^ The long-term impacts of this exposure on the natural history of hypertension remain unclear.

Lifecourse studies of in utero exposures and hypertension risk later in life are challenging. The latency period between exposure to the maternal in utero environment and onset of hypertension necessitates prolonged offspring follow-up with serial measures of confounding and possible mediating variables. A recent systematic review concluded that evidence of a direct effect of maternal pre-pregnancy body mass index (BMI) or weight, on offspring blood pressure later in life is lacking^11^. This uncertainty is due, in part, to the failure of previous studies to include potential mediators of this association, particularly growth trajectories through childhood and adolescence and to properly control for confounding.^11^ Obesity is an established risk factor for adolescent hypertension^12-14^, and weight trajectories throughout childhood and adolescence likely play an important role in mediating the association between maternal obesity and offspring hypertension in adolescence.

To our knowledge no study has incorporated serial measures of offspring weight trajectories in a causal analysis of the association between maternal pre-pregnancy weight status and offspring blood pressure later in life. The current study aimed to elucidate the impact of offspring growth trajectories on the development of elevated blood pressure in the presence of maternal weight status. We hypothesized that maternal pre-pregnancy BMI is associated with an increased risk of hypertension among offspring at 18 years of age, and a portion of the variance in this association is mediated indirectly through offspring weight trajectory (Figure 1).

**Figure 1.**
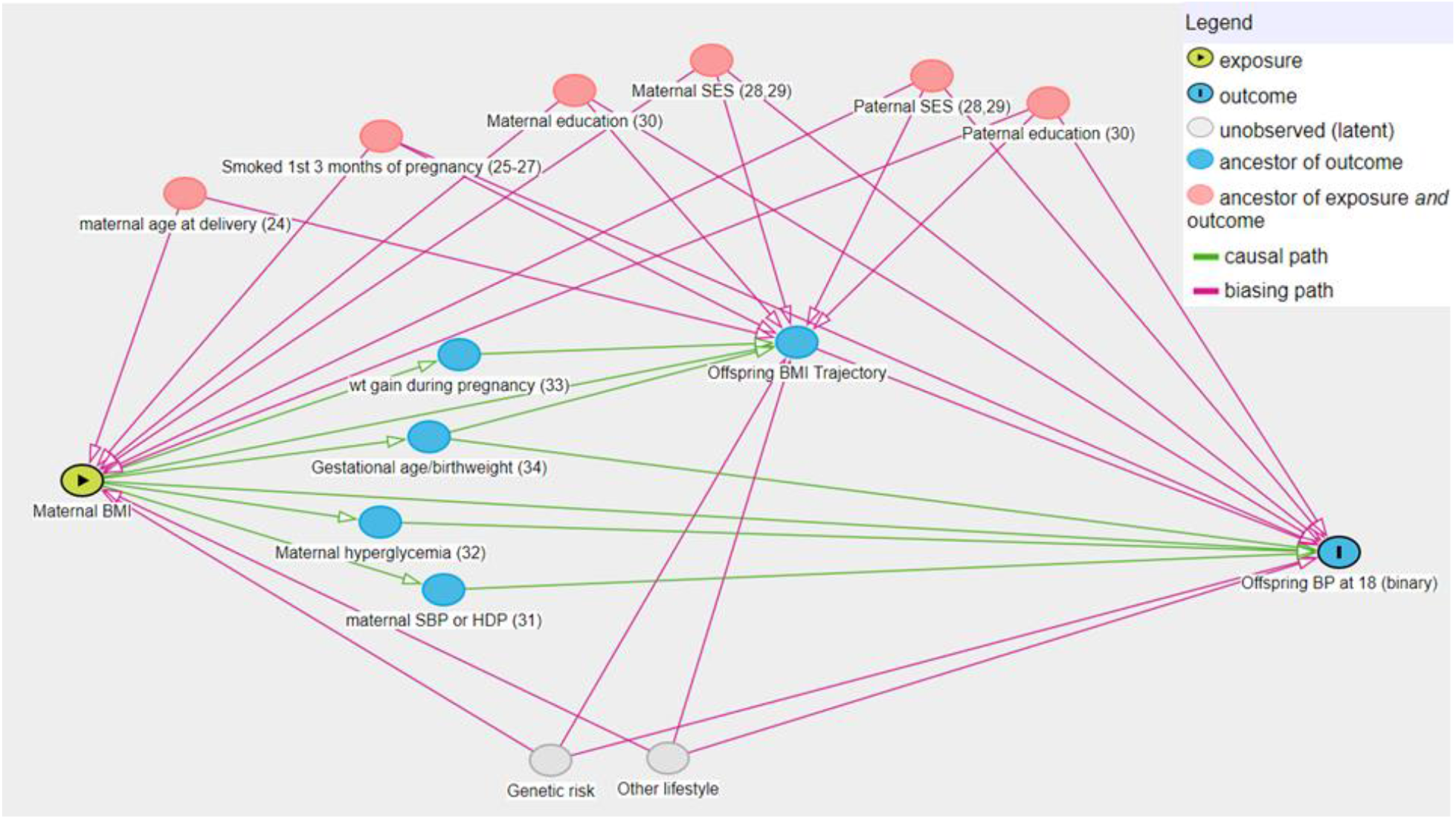
Directed acyclic graph (DAG)^46^ describing possible pathways through which exposure to maternal obesity may lead to elevated blood pressure in adolescence. BMI, body mass index; SES, socioeconomic status; SBP, systolic blood pressure; BP, blood pressure; HDP, hypertensive disorders of pregnancy. References in parentheses.

## METHODS

### Study Population

The Avon Longitudinal Study of Parents and Children (ALSPAC) cohort is a prospective population-based birth cohort study which originally recruited 14 541 pregnant women with delivery dates between April 1, 1991, and December 31, 1992. Of these initial pregnancies, there was a total of 14 062 live births and 13 988 children alive at 1 year of age. Beginning at 7 years of age (1998 to 1999), mothers and offspring were invited for annual or bi-annual follow-up clinic visits. Details of the cohort and the representativeness of the offspring are published.^15, 16^ the ALSPAC study website contains details of all the data that is available through a fully searchable data dictionary and variable search tool (http://www.bris.ac.uk/alspac/researchers/our-data). The present study used a subset of 3217 maternal-offspring pairs for whom pre-pregnancy height and weight measures were available and offspring attended all (n=7) clinic assessments between 7 and 18 years of age (April 1999 to December 31^st^ 2000).^17^ Ethical approval was obtained from the ALSPAC Law and Ethics Committee and the Biomedical Research Ethics Board at the University of Manitoba. Informed consent for the use of data collected via questionnaires and clinics was obtained from participants following thee recommendations of the ALSPAC Ethics and Law Committee at the time.

### Primary Exposure

The primary exposure of interest was maternal pre-pregnancy body mass index (BMI). Maternal pre-pregnancy weight and height were self-reported at the first clinical pregnancy visit and BMI was calculated as weight(kg)/height(m)^2^. Maternal BMI was treated as continuous in the causal mediation model. Maternal BMI was categorized for certain descriptive analyses according to World Health Organization categories (normal weight ≤ 24.9 kg/m^2^; overweight 25.0– 29.9 kg/m^2^; obese ≥ 30.0 kg/m^2^). Previous analyses from this cohort on systematic bias in self-report weight (e.g. heavier women may systematically under-reporting their weight) revealed that misreporting was similar for most participants and was not influenced by mean weight.^18^

### Outcome Variable of interest

The primary outcome, offspring blood pressure, was dichotomized as “normal” versus “elevated” according to the American Academy of Pediatrics 2017 guidelines for diagnosing elevated blood pressure and hypertension.^19, 20^ Offspring systolic and diastolic blood pressure were measured two or three times at the 18 – year follow up clinic with the participant at rest using a Dinamap 9301 Vital Signs Monitor.^15^ Arm circumference was measured, and an appropriate cuff size was chosen according to manufacturer’s instructions. The average of the final 2 readings was used in the analysis. Normal blood pressure was defined as systolic and diastolic blood pressure below 120 mmHg and 80 mmHg, respectively. Elevated blood pressure was defined as a systolic blood pressure above 120 mmHg or diastolic blood pressure above 80 mmHg to capture offspring with elevated blood pressure, stage 1, and stage 2 hypertension.

### Mediating Variable of interest

At each clinic visit from 7 to 18 years of age, offspring height and weight were measured using a standard protocol to the last complete millimetre using the Harpenden Stadiometer (Holtain Crosswell, Dyfed, UK) and to the nearest 50 g using the Tanita Body Fat Analyser (Model TBF 305, Tanita, Arlington Heights, IL), respectively. Raw weight and height measurements were converted to sex- and age-standardized BMI z-scores using the Canadian Pediatric Endocrine Group (CPEG) R Shiny app based on the World Health Organization growth charts (https://rpubs.com/AtulSharma/252678).

Offspring BMI z-scores were used in a data driven latent trajectory analysis (LTA) to quantify patterns of weight gain in offspring between 7 and 18 years of age. Latent class growth mixture models assume that there are multiple distinct trajectories each representing a hidden (i.e. latent) phenotypic class defined by its shape over time.^21^ Offspring with at least 2 data points for height and weight were used to model the distinct individual growth classes. With BMI z-score as the dependent variable, cubic models for age were fit with random intercepts and random linear slopes. Participant specific random effects allowed participants to vary around their class’ latent trajectory. The purpose of this analysis was to establish evidence for clusters and to concisely capture the essence of the multivariate BMI panel data for use as a mediator. The Bayesian Information Criterion (BIC) was used to enumerate latent classes for both boys and girls.^22^ Posterior class probabilities were used to evaluate model fit and assign offspring into their most likely latent growth class based on their observed BMI patterns. The latent class growth mixture models were fit using the lcmm package in R.^23^ For all analyses the mediator, offspring BMI class, was treated as a categorical variable.

### Descriptive and Confounding Variables

Maternal age at delivery,^24^ maternal smoking status during the first 3 months of pregnancy,^25-27^ maternal socioeconomic status (SES), paternal SES, maternal highest education attainment, and paternal highest education attainment^11, 28-30^ were identified as possible confounders (Figure 1) and adjusted for in all analyses. The above variables were chosen based on previous literature and availability in present dataset. Gestational weight gain, maternal systolic blood pressure or hypertensive disorders of pregnancy, hyperglycemia or diabetes in pregnancy, and offspring gestational age or birthweight have also been previously identified as having potential associations with offspring blood pressure at 18,^9, 31-33^ however these variables were not controlled for as they were seen as laying along the causal pathway of interest (Figure 1). Demographic variables were obtained from the obstetric records. Parental occupation was classified into social class groups from I (professional/managerial) to V (unskilled manual workers) using the 1991 British Office of Population and Census Statistics classification.^15^ Maternal and paternal highest education attainment were obtained from a questionnaire administered during pregnancy and used as secondary measures of social class.

## STATISTICAL ANALYSES

Normally distributed variables are expressed as mean ± SD. Comparisons among variables between offspring with elevated blood pressure and normal blood pressure were done using independent samples Student’s t-test or chi square test for categorical and binary variables. To robustly confirm that offspring BMI class was a possible mediating factor between maternal pre-pregnancy BMI and offspring blood pressure status, we conducted a series of multivariate regression analyses. Linear regression was used to assess mean differences in offspring blood pressure at 18 and maternal pre-pregnancy BMI among different offspring BMI classes. Multinomial logistic regression was used to assess associations between offspring BMI class and blood pressure status at 18 as well as maternal BMI categories and offspring BMI class.

Two distinct statistical approaches were used to assess the relationship between maternal pre-pregnancy BMI and offspring blood pressure status at 18 years. First, to initially test for an association, we assessed 3 multivariable logistic regression models: an unadjusted model (model 1), a model adjusted for potential confounders including maternal systolic blood pressure at 8 weeks gestation, maternal highest education attainment, and weeks gestation at delivery (model 2), and a model adjusting for the potential mediator, BMI class (model 3). The confounders in model 2 were identified based on previous literature as describe earlier.

The second approach used a counterfactual-based causal mediation analysis to assess the mediating effect of offspring BMI trajectory on the association between maternal BMI and offspring blood pressure status at 18. The goal of this analysis was to evaluate the potential mechanistic, or mediating, role of offspring BMI trajectory in the relationship between maternal BMI and offspring blood pressure at 18 years. Maternal BMI was left continuous for this analysis to capture all the information. The mediation analysis was completed using the Medflex package^34^ for R and the imputation method was used to estimate counterfactuals.^35, 36^ Medflex uses a natural effects model with nested counterfactuals to estimate the total effect of the exposure on the outcome and then decompose that into the natural direct effect (NDE) and the natural indirect effect (NIE). Simply put, to determine the NDE and NIE the mediator is held fixed and the exposure is allowed to vary (NDE); or the exposure is fixed and the mediator is allowed to change as though the exposure had changed (NIE) and the resulting change in the outcome is noted. The NDE and NIE are two causal components of the total effect of maternal pre-pregnancy BMI on offspring blood pressure status at 18. Further details on technical aspects of how the NDE and NIE are calculated and interpreted using the method described above are published elsewhere.^34, 37^ A p < 0.05 was considered statistically significant for groupwise comparisons and mediation analyses.

## RESULTS

Among the 14,855 women enrolled in the ALSPAC cohort, 3,217 mother-offspring pairs had valid data for maternal pre-pregnancy BMI, a minimum of 2 measures of offspring height and weight between 7 and 18 years of age, and offspring blood pressure at 18 years old. Compared to the full cohort, mothers included in analyses were older (29 vs 27 years, p < 0.01) and less likely to smoke (12 vs 22%, p < 0.01) on average compared to the rest of the cohort. The proportion of pregnancies complicated by pre-eclampsia, obesity, and hyperglycemia was similar between women included and those excluded from these analyses (appendix, Table S1). Offspring included in analyses were less likely to be born premature, had a higher mean birthweight, and at 7 and 18 years had slightly lower BMI, weight, and systolic blood pressure compared to the rest of the cohort (appendix, Table S1). Of the included offspring 55% were female, 676 (21%) were overweight or obese at 18 years of age, 865 (27%) were categorized as having elevated blood pressure, and 510 (16%) were considered hypertensive at 18 as per AAP cut-offs categories.

Baseline characteristics of mothers and offspring stratified according to offspring blood pressure status at 18 years of age are presented in Table 1. Maternal systolic blood pressure at 8 weeks was significantly higher among offspring with elevated blood pressure compared to those with normal blood pressure. There was also a higher proportion of caesarean sections among offspring with elevated blood pressure and a greater proportion of mothers without secondary or post secondary education qualifications. Weeks gestation at delivery was lower among offspring with elevated blood pressure whereas birthweight was similar between the two groups. Offspring with elevated blood pressure at 18 years of age had significantly higher systolic and diastolic blood pressure, body weight, and BMI at 7 and 18 years old compared to offspring with normal blood pressure. Participant characteristics stratified by maternal weight status (healthy weight, overweight, or obese) are presented in Table S2 of the appendix. Offspring born to mothers with pre-pregnancy obesity had higher systolic (121 vs 118 mmHg, p < 0.01) and diastolic blood pressure (66 vs 63 mmHg, p < 0.01) and BMI (34 vs 21 kg/m^2^, p < 0.01) at 18 years compared to offspring born to mothers with pre-pregnancy normal weight.

**Table 1.** Participant characteristics stratified according to offspring blood pressure status

|  | **Normal BP @ 18**<br>n = 1842 | **Elevated+ BP @ 18**<br>n = 1375 | **p value** |
| --- | --- | --- | --- |
| **Maternal Characteristics** |  |  |  |
| Age (yrs) | 39 ± 5 | 39 ± 5 | 0.81 |
| Parity | 1 ± 1 | 1 ± 1 | 0.67 |
| Gestation (weeks) | 39.6 ± 1.6 | 39.4 ± 1.8 | 0.001 |
| Pre-pregnancy BMI<br>(kg/m<sup>2</sup>) | 22.5 ± 3.4 | 23.2 ± 4.0 | < 0.001 |
| GWG (kg) | 12.7 ± 4.3 | 12.7 ± 4.4 | 0.98 |
| SBP at 8 weeks<br>(mmHg) | 112 ± 7 | 114 ± 7 | < 0.001 |
| Caesarean section (n/%) | 162 (14.4) | 153 (17.8) | 0.04 |
| Preeclampsia (n/%) | 5 (0.3) | 6 (0.4) | 0.54 |
| Hyperglycemia (n/%) | 82 (4.5) | 47 (3.4) | 0.17 |
| Smoke tobacco (n/%) | 208 (11.4) | 179 (13.1) | 0.15 |
| Social class (n/%) |  |  |  |
| *Manual labour*<br>*(semiskilled and*<br>*unskilled)* | 250 (15.4) | 171 (14.1) | 0.36 |
| Education (n/%) |  |  |  |
| *No education*<br>*qualification* | 166 (9.2) | 155 (11.4) | 0.04 |
| **Offspring Characteristics** |  |  |  |
| Birthweight (g) | 3448 ± 505 | 3429 ± 548 | 0.30 |
| Sex (F/M) | 1340/502 | 434/941 | < 0.001 |
| **At 7 years:** |  |  |  |
| DBP (mmHg) | 56 ± 6 | 57 ± 7 | < 0.001 |
| SBP (mmHg) | 96 ± 9 | 101 ± 9 | <0.001 |
| Weight (kg) | 24.8 ± 3.9 | 26.5 ± 4.5 | <0.001 |
| BMI (kg/m<sup>2</sup>) | 16 ± 2 | 16 ± 2 | <0.001 |
| **At 18 years:** |  |  |  |
| DBP (mmHg) | 62 ± 5 | 66 ± 7 | <0.001 |
| SBP (mmHg) | 111 ± 6 | 129 ± 7 | <0.001 |
| Weight (kg) | 62.1 ± 11.1 | 72.5 ± 14.0 | <0.001 |
| BMI (kg/m<sup>2</sup>) | 22 ± 3 | 24 ± 4 | <0.001 |

*mean ± SD. Social class: counts of manual labour workers (semiskilled and unskilled);*
*Education: counts of those with no education qualification; SBP: systolic blood pressure; DBP:*

The latent trajectory analysis yielded nearly identical growth classes for boys and girls (appendix, Figure S1) therefore data were pooled. Pooled data revealed 5 distinct trajectories which we labelled as (1) persistently elevated BMI Z (7.2%); (2) initial weight gain that quickly normalized (18.5%); (3) normal BMI Z throughout (43.7%); (4) initial weight gain that slowly normalized (n= 7.2%) and (5) persistently low BMI Z (n= 23.3%) (Figure 2). Offspring and maternal characteristics stratified by BMI Z class trajectory are presented in Table S3 of the appendix. Mean systolic blood pressure was higher from age 7 through to 18 among offspring in the persistently elevated BMI class compared to those in the normal BMI class and those in the persistently low BMI class had lower mean systolic blood pressure throughout childhood and adolescence (appendix, Figure S2). Multivariable logistic regression analyses revealed that the odds of elevated blood pressure at 18 was 3.08 (95% CI: 2.17-4.36) times higher among offspring in the persistently high BMI class compared to those with normal BMI trajectory (appendix, Table S4). Offspring from mothers with pre-pregnancy overweight and obesity were 2.67 (95% CI: 1.83-3.90) and 5.69 (95% CI: 3.42-9.48) times more likely to be in the persistently elevated BMI trajectory and 30 and 63% less likely to be in the persistently low BMI trajectory, compared to offspring born to mothers with pre-pregnancy healthy weight (appendix, Table S5). Collectively, these data reveal that BMI trajectories were associated with both offspring blood pressure and maternal pre-pregnancy BMI, supporting its role as a mediator of the association between these two variables.

**Figure 2.**
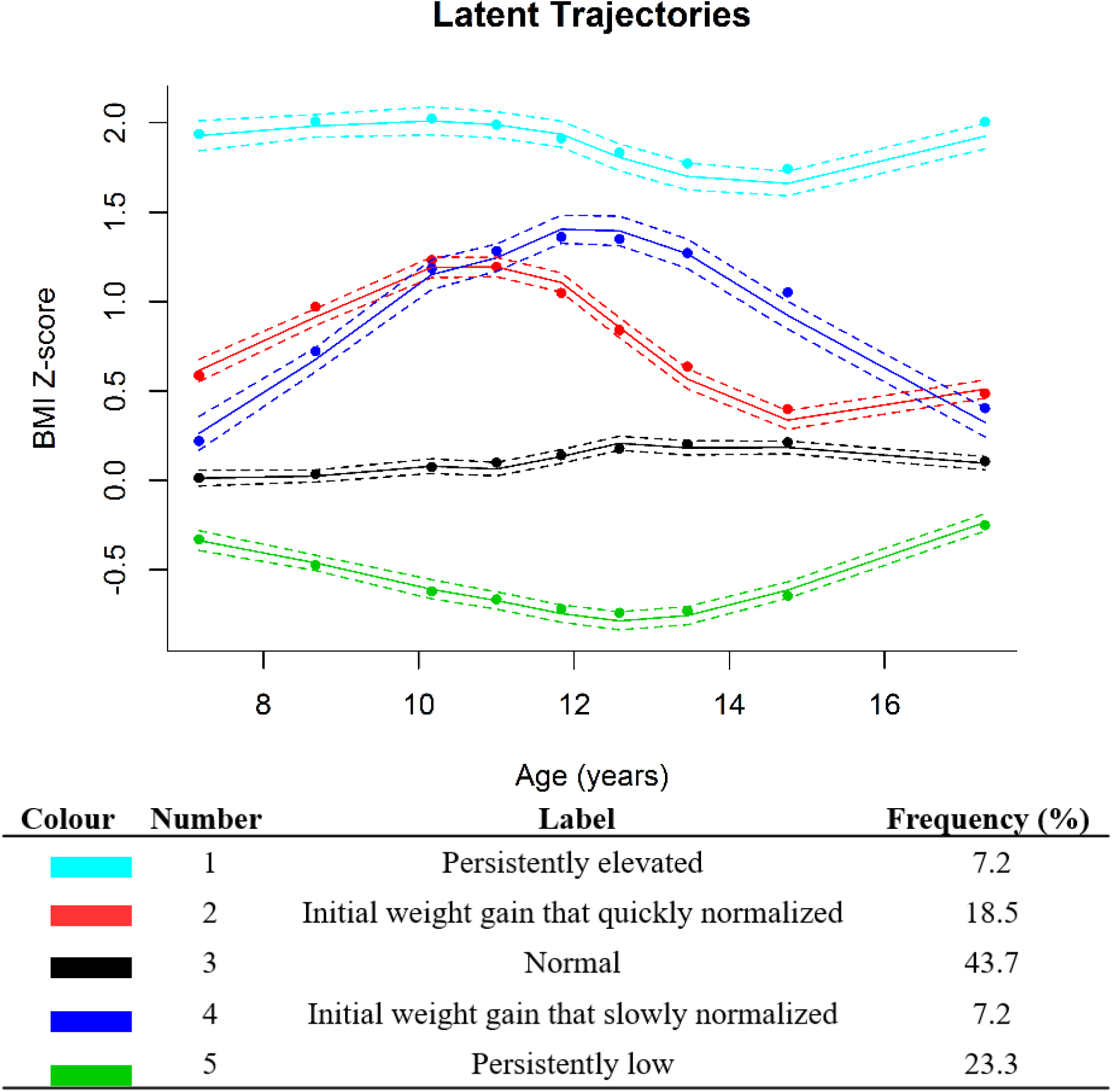
Offspring growth trajectories and descriptions standardized according to the world health organization. Pooled male and female data. BMI: body mass index

Results from multivariable logistic regression models are provided in Table 2. In unadjusted analyses, the odds of developing elevated blood pressure at 18 years of age was 5% higher for every 1 unit increase in maternal pre-pregnancy BMI (p< 0.001). After adjusting for relevant confounders, the risk of elevated blood pressure at 18 remained significant (aOR: 1.05, 95% CI: 1.03 – 1.07; p <0.001). Adding offspring BMI Z class trajectories in model 3 reduced the effect size where the risk of elevated blood pressure at 18 was only 3% higher per 1 unit increase in maternal pre-pregnancy BMI (aOR: 1.03; 95% CI: 1.00 – 1.05; p = 0.017). When sex was added to the model the strength of the association was no longer statistically significant (aOR: 1.02; 95% CI: 0.98 – 1.04; p = 0.08). In the final model (model 4) the odds of elevated blood pressure was 85% lower in girls compared to boys (aOR: 0.15; 95% CI: 0.12-0.18) and 4.5 fold higher (aOR: 4.5; 95% CI: 3.08-6.5) in offspring with a persistently elevated BMI Z score compared to those with a normal BMI Z score (Table 2). In sensitivity analyses, we did not observe an interaction between sex and maternal pre-pregnancy BMI.

**Table 2.** Associations between maternal pre-pregnancy BMI and elevated offspring SBP at 18

| | $\beta$ | OR | 95% Confidence Interval | P - value |
| --- | --- | --- | --- | --- |
| **Model 1** |  |  |  |  |
| Unadjusted | 0.048 | 1.05 | 1.03 – 1.07 | <0.001 |
| **Model 2** |  |  |  |  |
| Adjusted* | 0.046 | 1.05 | 1.03 – 1.07 | <0.001 |
| **Model 3** |  |  |  |  |
| Model 2 + BMIZ trajectory | 0.027 | 1.03 | 1.00 – 1.05 | 0.017 |
| **Model 4** |  |  |  |  |
| Model 3 + Sex | 0.022 | 1.02 | 0.98 – 1.05 | 0.08 |

\*adjusted for maternal and paternal SES, maternal and paternal highest education attainment, maternal age at delivery, and maternal smoking status in the first three months of pregnancy. BMIZ class: offspring body mass index z score trajectory based on latent class analysis. OR = Odds ratio

Results from the causal mediation analyses are shown in Figure 3. A natural effects model revealed that the total effect for a 5 unit increase in maternal BMI was a 22% increased risk of elevated blood pressure among offspring at 18 years old. The natural direct and natural indirect effects accounted for 54% and 46% of the total effect, respectively. Every 5 unit increase in maternal pre-pregnancy BMI directly increased offspring risk of elevated blood pressure at 18 by 13% (aOR = 1.12; 95% CI: 1.00-1.25; p = 0.04) and indirectly increased risk of elevated blood pressure at 18 by 10% (aOR = 1.1; 95% CI: 1.06, 1.14; p <0.001). Roughly half of the effect of maternal pre-pregnancy BMI was mediated by offspring BMI trajectory. Results were similar when stratified by sex (appendix, Figure S3); however, the direct effect was no longer significant in males (aOR 1.18; 95% CI: 0.95-1.31, p = 0.17) or females (aOR 1.16; 95% CI: 0.99-1.34, p = 0.06).

**Figure 3.**
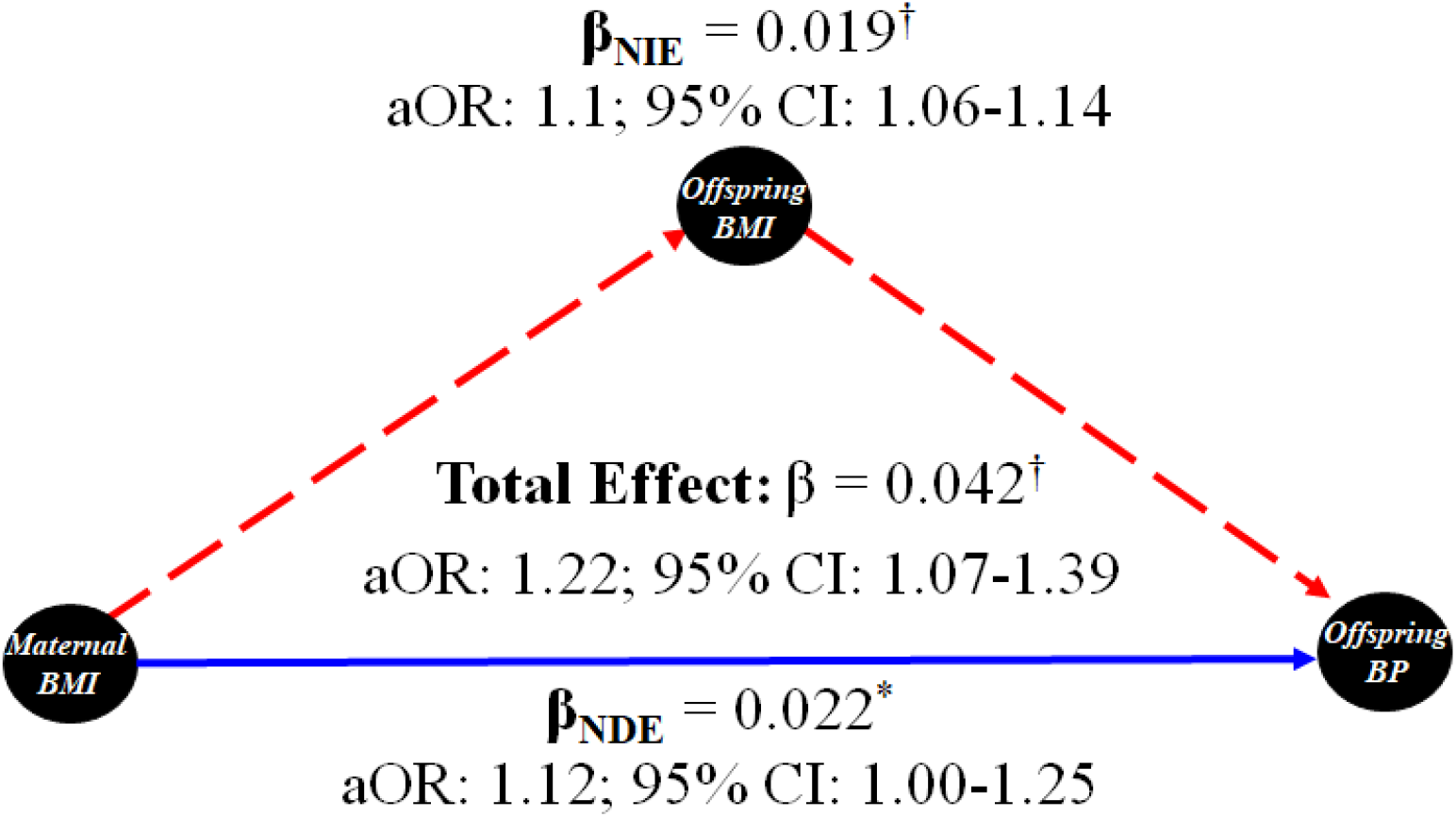
Simplified DAG showing results from causal mediation analysis. NDE, natural direct effect; NIE, natural indirect effect. Adjusted for maternal and paternal SES, maternal and paternal highest education attainment, maternal age at delivery, and maternal smoking status in the first three months of pregnancy. *p<0.05; †p < 0.0001. Adjusted odds ratios reflect a 5-unit increase in maternal body mass index.

## DISCUSSION

The series of analyses conducted within this large pregnancy cohort suggest that maternal pre-pregnancy obesity may act as a causal determinant of developing elevated blood pressure among offspring at 18 years of age. The relative proportion of offspring with elevated blood pressure reported here is similar to previous analyses performed with ALSPAC data.^38^ We identified a series of weight trajectories throughout mid-childhood and adolescence that mediate the effect of maternal BMI on offspring blood pressure status. Particularly, a sustained elevated BMI from 7 to 18 years of age is a robust mediator of being exposed to obesity in utero and elevated blood pressure risk at 18 years of age. These data provide robust evidence for direct and indirect effects for maternal pre-pregnancy obesity on the development of hypertension in late adolescence.

An emerging body of evidence suggests that exposure to an adverse metabolic milieu in utero is involved in the pathogenesis of early onset cardiovascular disease.^39^ The results from this study reinforce previous findings and provide insight into possible mechanisms. Several birth cohort studies (including the one used here) demonstrated that higher maternal pre-pregnancy BMI is positively associated elevated blood pressure at 4-6,^40^ 9^41^ and 15 years of age.^2^ Similar associations have been found with exposure to gestational diabetes in utero .^42^ The data presented here support these observations and extend them in several important ways. First, we provide more robust evidence that the association between elevated blood pressure in late adolescence is influenced by maternal pre-pregnancy BMI. Second, we demonstrated that this association is mediated through an elevated offspring BMI trajectory from childhood through adolescence. Lastly, in addition to the mediating effect of BMI trajectory, there is a direct effect of maternal pre-pregnancy BMI on adolescent blood pressure status at 18 years of age. Collectively, these data reinforce the concept that early life exposures, particularly maternal BMI, is involved in the natural history of adolescent hypertension directly and indirectly through offspring weight status.

Obesity is an established primary risk factor for hypertension in adolescence and persistent obesity throughout childhood and adolescence may confer additional risk.^14^ Previous pediatric cohort studies have documented four to five distinct weight trajectories defined by serial measures of BMI in both childhood and adolescence.^31, 43-45^ To the best of our knowledge, only two of these studies^14, 44^ used an unbiased data driven approach to determine these trajectories and none normalized BMI for age and sex. The data presented here, using latent class growth mixture modeling revealed five distinct trajectories of BMI z – scores throughout childhood and adolescence. Like previous cohort studies, we found that three trajectories were associated with an obesity phenotype; two that involved rapid weight gain in childhood and early adolescence which normalized in late adolescence, and one that involved persistent obesity throughout. Persistent obesity was robustly associated with higher blood pressure throughout adolescence and a substantially increased odds of elevated blood pressure and hypertension at 18 years of age. A novel addition to previous literature was the observation that persistently high BMI from 7 to 18 years of age was also associated with maternal pre-pregnancy obesity, independent of measured confounding (appendix, Table S5). Additionally, BMI trajectory from 7 to 18 years of age was a significant mediator of the association between maternal pre-pregnancy BMI and blood pressure at 18 years of age. These data reinforce the concept that BMI throughout childhood and adolescence may mediate associations between maternal exposures in pregnancy and risk for early-onset cardiovascular disease.

The study presented here is strengthened using a highly phenotyped large sample of women and their offspring that provided serial measurements of mediators and outcomes for nearly two decades. Despite these strengths, the study suffers from several limitations that need to be addressed. As this was an observational study there is a risk that some degree of unmeasured confounding could have influenced these results. An important assumption of the causal mediation analysis described above states that there is no unmeasured confounding between the exposure, mediator, and the outcome, however there is likely some residual confounding in the present model (Figure 1). The most important confounder being genetic risk of elevated blood pressure among both mother and offspring. Including genetic information in the analyses will be an important next step moving forward. Potentially important lifestyle factors like physical activity and diet were also not controlled for here. Finally, measures of offspring BMI before 7 years of age were not captured in the ALSPAC cohort and thus were not included in the latent class analysis which limited our ability to assess growth trajectories in early childhood.

Despite the important limitations described above, the present study demonstrates that maternal BMI prior to pregnancy is positively, though modestly, associated with elevated offspring blood pressure at 18 years of age. Maternal pre-pregnancy BMI has a nearly equal direct positive effect on offspring blood pressure at 18 years of age and an indirect effect mediated by offspring BMI trajectory. These data support the concept that early life exposures, particularly maternal obesity, may play a role in the development of adolescent elevated blood pressure and that interventions targeted at lowering offspring BMI may have a protective effect against elevated offspring blood pressure at 18 years of age, despite being exposed to overweight or obesity in utero.

## Data Availability

All data were purchased from the Avon Longitudinal Study of Parents and Children (ALSPAC) cohort.

http://www.bristol.ac.uk/alspac/researchers/access/

## Acknowledgements

We are extremely grateful to all the families who took part in the ALSPAC study, the midwives for their help in recruiting them, and the whole ALSPAC team, which includes interviewers, computer and laboratory technicians, clerical workers, research scientists, volunteers, managers, receptionists, and nurses.

## Author contributions

NB, BD and JMc

MA

AD

## Notes

### Competing Interest Statement

The authors have declared no competing interest.

### Funding Statement

The UK Medical Research Council and Wellcome (Grant ref: 217065/Z/19/Z) and the University of Bristol provide core support for ALSPAC. A comprehensive list of grants funding is available on the ALSPAC website (http://www.bristol.ac.uk/alspac/external/documents/grant-acknowledgements.pdf). JMM was supported by a Canadian Institutes of Health Research (CIHR) Applied Public Health Chair, MBA holds a Tier 2 Canada Research Chair (CRC) in the Developmental Origins of Chronic Disease.
JMM holds grants from the CIHR, Diabetes Canada, the Heart and Stroke Foundation (HSF), the Cosmopolitan Foundation, and the Lawson Foundation. AD currently has research funding from CIHR, Research Manitoba and CHRIM. MBA holds grants from the CIHR, The Bill and Melinda Gates Foundation, the Canada Foundation for Innovation, the Garfield G. Weston Foundation, Prolacta Biosciences, Mitacs, the Canadian Institutes for Advanced Research, the Childrens Hospital Foundation of Manitoba, and Research Manitoba; NB and Drs JMM, MBA and AD were also supported by a Research Manitoba grant to the DEVOTION research cluster. None of the cited agencies were involved in the design, conduct, or approval of this manuscript.

### Author Declarations

The University of Manitoba Health Research Ethics Board file number HS22554(H2019:055).

