## Supplemental material for "Maternal obesity before pregnancy predicts offspring blood pressure at 18 years of age: A causal mediation analysis"

### Appendix: Supplementary tables and figures

**Table S1.** Differences in demographics between the entire ALSPAC cohort and the subset included in our mediation analysis

| | Whole cohort (n = 11,638) | Causal mediation cohort (n=3217) | p value |
|---|---|---|---|
| **Maternal Characteristics** | | | |
| Maternal age | 27.5 ± 5.00 | 29.5 ± 4.56 | < 0.001 |
| Pre-pregnancy BMI | 23.0 ± 3.92 | 22.8 ± 3.69 | 0.05 |
| Weight gain in pregnancy | 12.5 ± 4.72 | 12.7 ± 4.35 | 0.16 |
| SBP at 8 weeks | 113 ± 7 | 113 ± 7 | 0.55 |
| Pre-eclampsia | 20 (0.19) | 11 (0.34) | 0.17 |
| Hyperglycemia in pregnancy | 379 (4.2) | 129 (4.0) | 0.76 |
| Social class | 2307 (31) | 417 (22) | < 0.001 |
| **Offspring Characteristics** | | | |
| Sex (female) | 5489 (47.2) | 1774 (55.1) | < 0.001 |
| Pre-term birth | 1308 (12.2) | 295 (9.2) | < 0.001 |
| Birthweight | 3379 ± 583 | 3440 ± 524 | < 0.001 |
| BMI at 7 (kg/m$^2$) | 16.4 ± 2.2 | 16.1 ± 1.9 | < 0.001 |
| Weight at 7 (kg) | 26.2 ± 5.0 | 25.5 ± 4.2 | < 0.001 |
| SBP at 7 (mmHg) | 99.3 ± 9.3 | 98.5 ± 9.0 | < 0.001 |
| DBP at 7 (mmHg) | 56.6 ± 6.7 | 56.4 ± 6.6 | 0.30 |
| BMI at 18 (kg/m$^2$) | 23.3 ± 4.5 | 22.6 ± 4.0 | < 0.001 |
| Weight at 18 (kg) | 68.3 ± 14.9 | 66.5 ± 13.4 | < 0.001 |
| SBP at 18 (mmHg) | 119 ± 11 | 119 ± 11 | 0.84 |
| DBP at 18 (kg/m$^2$) | 65 ± 7 | 64 ± 7 | < 0.001 |

*Mean ± SD or n (%); Social class: manual labour (partly skilled, or unskilled); BMI: body mass index; SBP: systolic blood pressure; DBP: diastolic blood pressure*

**Table S2**. Baseline demographics stratified by pre-pregnancy weight status

| Variables | Healthy Weight (n = 2568) | Overweight (n = 489) | Obese (n = 160) |
|---|---|---|---|
| **Maternal Characteristics** | | | |
| Pre pregnancy BMI | 21 ± 2 | 27 ± 1** | 34 ± 4ǂ |
| Age | 29.6 ± 4.5 | 29.4 ± 4.8 | 28.9 ± 5.1 |
| SBP at 8 weeks | 112 ± 7 | 115 ± 7** | 119 ± 8ǂ |
| DBP at 8 weeks | 66 ± 4 | 68 ± 4** | 71 ± 5ǂ |
| Pregnancy weight gain (kg) | 12.8 ± 4.1 | 12.8 ± 5.0 | 10.6 ± 5.6ǂ |
| Preeclampsia | 6 (0.2) | 2 (0.4) | 3 (1.9)* |
| HPG | 87 (3.4) | 25 (5.1) | 17 (10.7)** |
| Tobacco use | 301 (11.8) | 65 (13.4) | 21 (13.1) |
| Low social class | 187 (9) | 40 (10) | 20 (17)* |
| **Offspring Characteristics** | | | |
| Female | 1421 (55) | 265 (54) | 88 (55) |
| Preterm birth | 248 (9.7) | 35 (7.2) | 12 (7.5) |
| Birthweight (g) | 3404 ± 514 | 3558 ± 518** | 3644 ± 599** |
| **At 18 years:** | | | |
| SBP (mmHg) | 118 ± 11 | 120 ± 11* | 121 ± 12** |
| DBP (mmHg) | 63 ± 6 | 64 ± 7* | 66 ± 7ǂ |
| Weight (kg) | 64.9 ± 12.0 | 71.0 ± 15.1** | 78.1 ± 20.0ǂ |
| BMI ($kg/m^2$) | 22 ± 3 | 24 ± 4** | 27 ± 6ǂ |

*Mean ± SD or n (%); *significantly different from normal weight at p <0.05; **significantly different from normal weight at p < 0.001; ǂsignificantly different from overweight; Low social class: manual labour (semiskilled and unskilled); SBP: systolic blood pressure; DBP: diastolic blood pressure; BMI: body mass index; HPG: hyperglycemia in pregnancy*

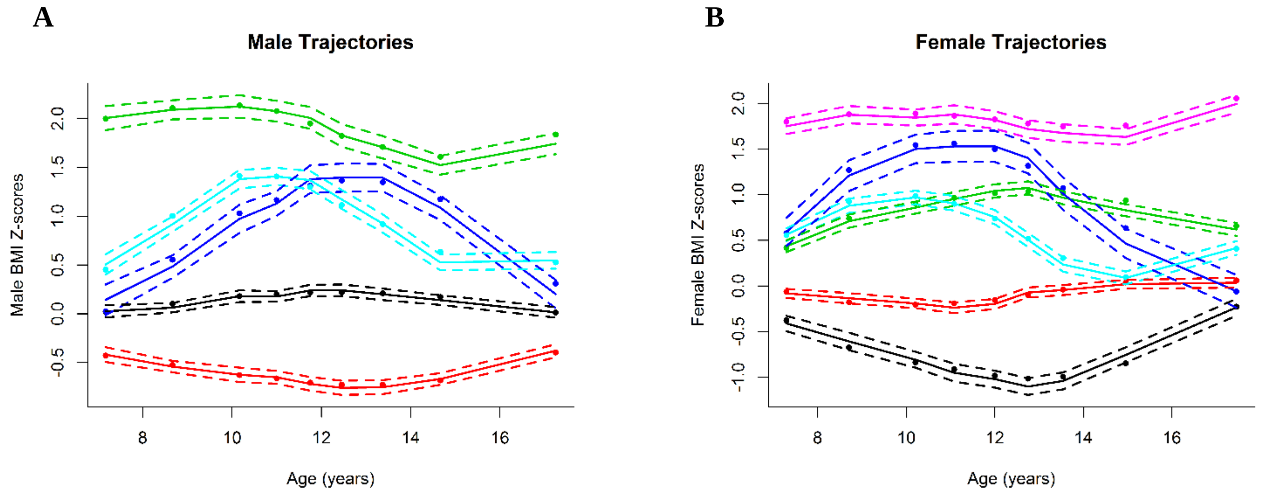


**Figure S1**. Offspring growth trajectories identified by latent trajectory analysis and stratified by sex. Colours in these figures are arbitrary and are simply to distinguish between different trajectories. (A) male trajectories and (B) female trajectories.

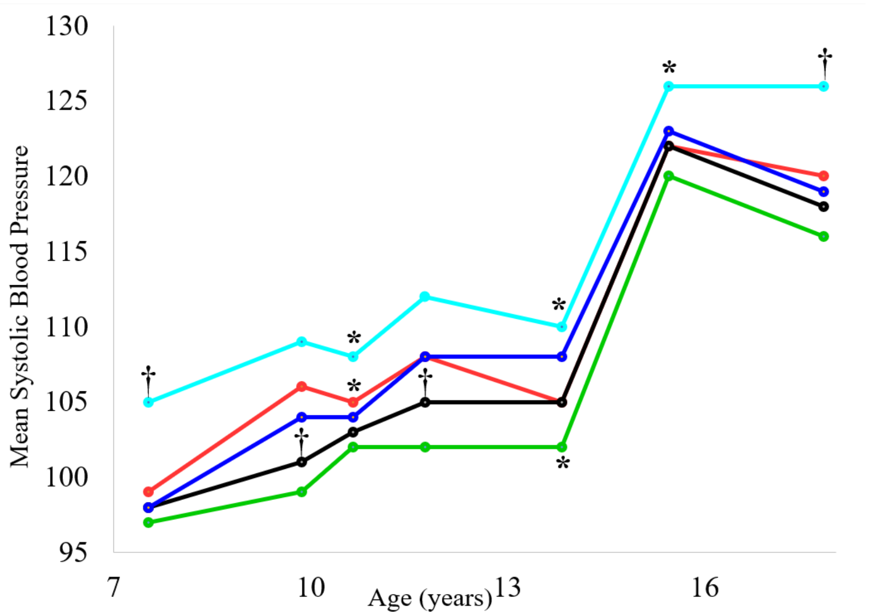


**Figure S2.** Mean systolic blood pressure from 7 to 18 years stratified by class. Colour coded to match BMIT trajectories: Light blue, persistently elevated BMI; red, initial weight gain that quickly normalized; black, normal; dark blue, initial weight gain that slowly normalized; green, persistently low BMI. *****significantly different from reference (black) at $p < 0.001$. †significantly different from all other classes at $p < 0.001$.

**Table S3**. Maternal and offspring characteristics stratified by offspring BMI trajectory

| | Persistently elevated *N = 233 (7.2%)* | Initial weight gain that quickly normalized *N = 595 (18.5%)* | Normal *N = 1406 (43.7%)* | Initial weight gain that slowly normalized *N = 233 (7.2%)* | Persistently low *N = 750 (23.3%)* | P value |
|---|---|---|---|---|---|---|
| **Maternal Characteristics** | | | | | | |
| pre-pregnancy BMI | 25.8 ± 5.3 | 23.1 ± 3.4 | 22.6 ± 3.4 | 23.9 ± 4.3 | 21.8 ± 2.9 | < 0.001 |
| Age in pregnancy | 29 ± 5 | 30 ± 5 | 29 ± 4 | 30 ± 5 | 30 ± 5 | 0.13 |
| Pre-eclampsia | 2 (0.9) | 1 (0.2) | 5 (0.4) | 0 (0) | 3 (0.4) | 0.54 |
| Hyperglycemia in pregnancy | 15 (6.5) | 21 (3.6) | 52 (3.7) | 11 (4.7) | 30 (4.0) | 0.33 |
| Smoking | 36 (15.5) | 72 (12.2) | 17 (12.6) | 28 (12.1) | 76 (10.2) | 0.25 |
| Social class | 22 (11) | 40 (7.5) | 103 (8.3) | 22 (10.9) | 60 (9.0) | 0.43 |
| **Offspring Characteristics** | | | | | | |
| Birthweight (g) | 3563 ± 487 | 3455 ± 508 | 3422 ± 516 | 3486 ± 583 | 3409 ± 537 | < 0.001 |
| Gestation (weeks) | 39.7 ± 1.5 | 39.6 ± 1.5 | 39.5 ± 1.7 | 39.4 ± 2.2 | 39.5 ± 1.8 | 0.33 |
| Sex (female) | 138 (59.2) | 320 (53.8) | 765 (54.4) | 96 (41.2) | 455 (60.7) | < 0.001 |
| **At 7 years** | | | | | | |
| BMI ($kg/m^2$) | 20.2 ± 2.3 | 16.8 ± 1.7 | 15.7 ± 1.3 | 16.2 ± 1.5 | 15.1 ± 1.3 | < 0.001 |
| SBP (mmHg) | 105 ± 10 | 99 ± 9 | 98 ± 9 | 98 ± 9 | 97 ± 9 | < 0.001 |
| DBP (mmHg) | 59 ± 7 | 57 ± 7 | 56 ± 6 | 56 ± 6 | 56 ± 6 | < 0.001 |
| **At 13 years** | | | | | | |
| BMI ($kg/m^2$) | 25.9 ± 3.8 | 20.8 ± 2.7 | 20.0 ± 2.4 | 23.4 ± 3.0 | 17.5 ± 1.7 | < 0.001 |
| SBP (mmHg) | 110 ± 11 | 105 ± 10 | 105 ± 9 | 108 ± 10 | 102 ± 8 | < 0.001 |
| DBP (mmHg) | 59 ± 7 | 57 ± 6 | 57 ± 7 | 57 ± 6 | 56 ± 6 | < 0.001 |
| **At 18 years** | | | | | | |
| BMI ($kg/m^2$) | 30.9 ± 4.8 | 23.4 ± 3.2 | 21.9 ± 2.9 | 22.7 ± 3.4 | 20.8 ± 3.0 | < 0.001 |
| SBP (mmHg) | 126 ± 12 | 120 ± 11 | 118 ± 10 | 119 ± 12 | 116 ± 10 | < 0.001 |
| DBP (mmHg) | 69 ± 8 | 64 ± 6 | 63 ± 6 | 63 ± 6 | 62 ± 6 | < 0.001 |

*Mean ± SD or n (%). Social class: manual labour (semiskilled and unskilled); BMI: body mass index; SBP: systolic blood pressure; DBP: diastolic blood pressure*

**Table S4.** Offspring BMI Z score trajectory is associated with blood pressure at 18 years of age.

| | **Mean difference in SBP at 18 (95% CI)** | **OR of SBP categories (95% CI)** | |
|---|---|---|---|
| | | **Elevated +** | **Normal** |
| Persistently elevated | 7.81 (6.17, 9.46) | 3.08 (2.17, 4.36) | 1.00 |
| Initial weight gain that quickly normalized | 1.79 (0.68, 2.9) | 1.38 (1.04, 1.82) | 1.00 |
| Normal | 0 | 1.00 | 1.00 |
| Initial weight gain that slowly normalized | 0.95 (0.-0.68, 2.58) | 1.11 (0.72, 1.70) | 1.00 |
| Persistently low | -1.45 (-2.47, -0.44) | 0.75 (0.56, 1.01) | 1.00 |

*Adjusted for maternal and paternal SES, maternal and paternal highest education attainment, maternal age at delivery, and maternal smoking status in the first three months of pregnancy; SBP: systolic blood pressure.*

**Table S5**. Maternal pre-pregnancy BMI is associated with offspring BMIZ trajectory

| | **Persistently elevated** | **Initial weight gain that quickly normalized** | **Normal** | **Initial weight gain that slowly normalized** | **Persistently low** |
|---|---|---|---|---|---|
| Per 1-unit maternal BMI (kg/m2) | 1.17 (1.13, 1.21) | 1.05 (1.02, 1.08) | 0 | 1.10 (1.06, 1.15) | 0.92 (0.89, 0.95) |
| BMI Categories: | | | | | |
| Normal | 1.00 | 1.00 | 1.00 | 1.00 | 1.00 |
| Overweight | 2.67 (1.83, 3.90) | 1.36 (1.03,1.81) | 1.00 | 1.33 (0.87,2.04) | 0.70 (0.52,0.95) |
| Obese | 5.69 (3.42, 9.48) | 1.22 (0.72, 2.06) | 1.00 | 3.25 (1.86,5.68) | 0.37 (0.19,0.75) |

*Adjusted for maternal and paternal SES, maternal and paternal highest education attainment, maternal age at delivery, and maternal smoking status in the first three months of pregnancy; BMI: body mass index.*

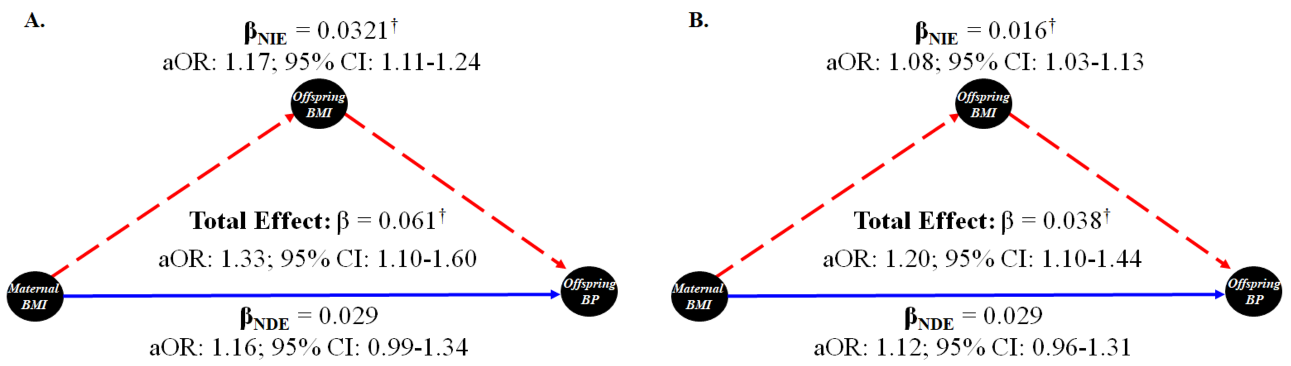


**Figure S3.** Simplified DAG showing results from causal mediation analysis stratified by sex: (A) Female; (B) Male. NDE, natural direct effect; NIE, natural indirect effect. Adjusted for maternal and paternal SES, maternal and paternal highest education attainment, maternal age at delivery, and maternal smoking status in the first three months of pregnancy. *$p<0.05$; †$p < 0.0001$. Adjusted odds ratios reflect a 5-unit increase in maternal body mass index.
